# Changes in English medication safety indicators throughout the COVID-19 pandemic: a federated analysis of 57 million patients’ primary care records in situ using OpenSAFELY

**DOI:** 10.1101/2022.05.05.22273234

**Authors:** Louis Fisher, Lisa E. M. Hopcroft, Sarah Rodgers, James Barrett, Kerry Oliver, Anthony J. Avery, Dai Evans, Helen Curtis, Richard Croker, Orla Macdonald, Jessica Morley, Amir Mehrkar, Seb Bacon, Simon Davy, Iain Dillingham, David Evans, George Hickman, Peter Inglesby, Caroline E. Morton, Becky Smith, Tom Ward, William Hulme, Amelia Green, Jon Massey, Alex J. Walker, Chris Bates, Jonathan Cockburn, John Parry, Frank Hester, Sam Harper, Shaun O’Hanlon, Alex Eavis, Richard Jarvis, Dima Avramov, Paul Griffiths, Aaron Fowles, Nasreen Parkes, Ben Goldacre, Brian MacKenna

## Abstract

**Objective:** To describe the impact of the COVID-19 pandemic on safe prescribing, using the PINCER prescribing indicators; to implement complex prescribing indicators at national scale using GP data.

**Design:** Population based cohort study, with the approval of NHS England using the OpenSAFELY platform.

**Setting:** Electronic health record data from 56.8 million NHS patients’ general practice records.

**Participants:** All NHS patients registered at a GP practice using TPP or EMIS computer systems and recorded as at risk of at least one potentially hazardous PINCER indicator between September 2019 and September 2021.

**Main outcome measure:** Monthly trends and between-practice variation for compliance with 13 PINCER measures between September 2019 and September 2021.

**Results:** The indicators were successfully implemented across GP data in OpenSAFELY. Hazardous prescribing remained largely unchanged during the COVID-19 pandemic, with only small reductions in achievement of the PINCER indicators. There were transient delays in blood test monitoring for some medications, particularly ACE inhibitors. All indicators exhibited substantial recovery by September 2021. We identified 1,813,058 patients at risk of at least one hazardous prescribing event.

**Conclusion:** Good performance was maintained during the COVID-19 pandemic across a diverse range of widely evaluated measures of safe prescribing.

**Summary box:** WHAT IS ALREADY KNOWN ON THIS TOPIC

- Primary care services were substantially disrupted by the COVID-19 pandemic.
- Disruption to safe prescribing during the pandemic has not previously been evaluated.
- PINCER is a nationally adopted programme of activities that aims to identify and correct hazardous prescribing in GP practices, by conducting manual audit on subgroups of practices.

WHAT THIS STUDY ADDS

- For the first time, we were able to successfully generate data on PINCER indicators for almost the whole population of England, in a single analysis.
- Our study is the most comprehensive assessment of medication safety during the COVID-19 pandemic in England, covering 95% of the population using well-validated measures.
- Good performance was maintained across many PINCER indicators throughout the pandemic.
- Delays in delivering some medication-related blood test monitoring were evident though considerable recovery was made by the end of the study period.

## Background

The World Health Organisation (WHO) launched a patient safety challenge in 2017, Medication Without Harm (1), with an ambition to “*reduce severe avoidable medication related harm globally by 50% in the next 5 years*” (2). The COVID-19 pandemic has affected the delivery of primary care services within the NHS (3–5). It is possible that this disruption will have contributed towards increased rates of medication related harm, with 34% of an estimated 66 million potentially clinically significant errors occurring in primary care prescribing in England annually as estimated using NHS dispensing statistics in 2015-16 (6).

As part of its response to the WHO challenge, PRIMIS at the University of Nottingham led on the national rollout of PINCER in collaboration with the NHS Academic Health Science Networks (7). The PINCER intervention (Box 1) is a proven programme of activities for reducing hazardous prescribing in general practices (8). Briefly, it involves training pharmacists to provide feedback, educational outreach and dedicated support to general practices, systematically focusing on patients identified to be at risk of harm from medications. These patients are identified using pre-specified and quality assured analytic indicators in the SNOMED-CT code classification system used by GP systems in England. PINCER includes 13 indicators of hazardous prescribing of high risk medications that: (1) can cause gastrointestinal (GI) bleeds; (2) are cautioned against in certain conditions (specifically heart failure, asthma and chronic renal failure); (3) require blood test monitoring. These indicators have been developed in collaboration with academics from the University of Nottingham and made available to pharmacists in practices participating in the PINCER programme.

OpenSAFELY is a new secure analytics platform for electronic patient records built by our group with the approval of NHS England to deliver urgent academic (9) and operational NHS service research (10,11) on the direct and indirect impacts of the pandemic: analyses can currently run across patients’ full raw pseudonymised primary care records at 95% of English general practices (55% in practices using EMIS software, and 40% in practices using TPP software) with patient-level linkage to various sources of secondary care data; all code and analysis is shared openly for inspection and re-use. The PINCER indicators created by PRIMIS are typically implemented for single practices, or groups of practices, through a variety of technical methods in different settings, to monitor compliance for practices that are participating in the PINCER programme.

We set out to implement the full suite of PINCER codelists, methods and indicators in OpenSAFELY to permit monitoring of compliance on all prescribing safety indicators at a population-level; and to describe changes in compliance during the COVID-19 pandemic.

### BOX 1

**PINCER BACKGROUND**

The PINCER intervention was developed and tested by researchers at the Universities of Nottingham, Manchester and Edinburgh and comprises three core elements (12):

1. Searching GP computer systems to identify patients at risk of potentially hazardous prescribing using a set of prescribing safety indicators. PINCER indicators are typically generated by conducting individual audits on subgroups of practices.
2. Pharmacists, specifically trained to deliver the intervention, providing an educational outreach intervention where they meet with GPs and other practice staff to:
  a. Discuss the search results and highlight the importance of the hazardous prescribing identified using brief educational materials
  b. Agree an action plan for reviewing patients identified as high risk and improving prescribing and medication monitoring systems using root cause analysis (RCA) to minimise future risk
3. Pharmacists (and pharmacy technicians) working with, and supporting, general practice staff to implement the agreed action plan.

Findings from the PINCER trial, published in the Lancet (8), demonstrated that PINCER is an effective and cost-effective method for reducing a range of clinically important and commonly made medication errors in primary care. For example, at 6 months’ follow-up, patients in the PINCER group were significantly less likely to have been prescribed an oral non-steroidal anti-inflammatory drug (NSAID) if they had a history of peptic ulcer without gastroprotection (Odds ratio (OR): 0·58; 95% Confidence Interval (CI): 0·38–0·89), thereby reducing their risk of hospital admission with gastrointestinal (GI) bleeding.

Over the last three years, PRIMIS—a unit of the University of Nottingham providing expert advice on the intelligent use of primary care data to the NHS, academics, and industry partners—has led on the national rollout of PINCER in collaboration with the Academic Health Science Network (7).

## Methods

### Study design

We conducted a retrospective cohort study using general practice primary care electronic health record (EHR) data from all GP practices in England supplied by the EHR vendors TPP and EMIS.

### Data source

Primary care records managed by the GP software providers TPP and EMIS are available in OpenSAFELY, a data analytics platform created by our team with the approval of NHS England to address urgent COVID-19 research questions (https://opensafely.org). OpenSAFELY provides a secure software interface allowing the analysis of pseudonymized primary care patient records from England in near real-time within the EHR vendor’s highly secure data centre, avoiding the need for large volumes of potentially disclosive pseudonymized patient data to be transferred off-site. This, in addition to other technical and organisational controls, minimises any risk of re-identification. Similarly pseudonymized datasets from other data providers are securely provided to the EHR vendor and linked to the primary care data. The TPP dataset analysed within OpenSAFELY (hereafter referred to as OpenSAFELY-TPP) is based on 24.2 million people currently registered with 2546 GP surgeries using TPP SystmOne software; the EMIS dataset analysed within OpenSAFELY (hereafter referred to as OpenSAFELY-EMIS) is based on 32.6 million people currently registered with 3821 GP surgeries using EMIS. It includes pseudonymized data such as coded diagnoses, medications and physiological parameters. No free text data are included. Further details on our information governance can be found under information governance and ethics.

### Study population

We included all patients who were: alive; aged 18-120; registered with an OpenSAFELY-TPP or OpenSAFELY-EMIS practice; and recorded as at risk of at least one potentially hazardous prescribing indicator as defined by the SNOMED-CT and NHS dictionary of medicines and devices (dm+d) codes developed by PRIMIS for the PINCER programme (https://www.opencodelists.org/codelist/pincer/) between September 2019 and September 2021 to adequately cover both the period of onset and subsequent recovery of the COVID-19 pandemic.

### Study measures

Definitions of the hazardous prescribing indicators are provided in Table 1. The percentage for each indicator is formed of a numerator and denominator pair, where the numerator is the set of patients deemed by the indicator to be at risk of a potentially hazardous prescribing event, and the denominator is the set of patients for which assessment of the indicator is clinically meaningful. Higher indicator percentages represent potentially poorer performance on medication safety. Indicators belong to three groups: (1) those associated with gastrointestinal bleeds; (2) those associated with cautioned medications; and (3) those associated with blood test monitoring; these groups are used to summarise results.

**Table 1.** PINCER indicator definitions.

| Description | Denominator | Numerator (group at risk to the potentially hazardous prescribing event) |
| --- | --- | --- |
| **Indicators associated with gastrointestinal bleeding** |  |  |
| **Age ≥ 65 & NSAID:** prescription of an oral non-steroidal anti-inflammatory drug (NSAID), without co-prescription of an ulcer-healing drug, to a patient aged ≥65 years | Patients aged ≥65 years without co-prescription of an ulcer-healing drug (proton pump inhibitor (PPI) or H2 antagonist) in the 3 months leading up to the audit date | Patients in the group denominator AND prescribed an oral NSAID in the 3 months leading up to the audit date |
| **PU & NSAID:** prescription of an oral NSAID, without co-prescription of an ulcer healing drug, to a patient with a history of peptic ulceration | Patients aged ≥18 years with a Read code for peptic ulcer or GI bleed at least 3 months before audit date and not prescribed an ulcer healing drug (PPI or H2 antagonist) within the 3 months leading up to the audit date | Patients in the group denominator AND prescribed an oral NSAID within the 3 months leading up to the audit date |
| **PU & antiplatelet:** prescription of an antiplatelet drug without co-prescription of an ulcer-healing drug, to a patient with a history of peptic ulceration | Patients aged ≥18 years with a Read code for peptic ulcer or GI bleed at least 3 months before audit date and not prescribed an ulcer healing drug (PPI or H2 antagonist) within the 3 months leading up to the audit date | Patients in the group denominator AND prescribed an antiplatelet drug (aspirin or clopidogrel or prasugrel or ticagrelor) within the 3 months leading up to the audit date |
| **Warfarin/DOAC & NSAID:** prescription of warfarin or DOAC in combination with an oral NSAID | Patients aged ≥18 years prescribed warfarin or a DOAC (apixaban or dabigatran or rivaroxaban or edoxaban) within the 3 months leading up to the audit date | Patients in the group denominator AND prescribed an oral NSAID within the 3 months leading up to the audit date |
| **Warfarin/DOAC & antiplatelet:** prescription of warfarin or DOAC and an antiplatelet drug in combination without coprescription of an ulcer-healing drug | Patients aged ≥18 years prescribed warfarin or DOAC without co-prescription of ulcer-healing drug (PPI or H2 antagonist) within the 3 months leading up to the audit date | Patients in the group denominator AND prescribed an antiplatelet drug (aspirin or clopidogrel or prasugrel or ticagrelor) within the 3 months leading up to the audit date and within 28 days of the warfarin/ DOAC prescription |
| **Aspirin & other antiplatelet:** prescription of aspirin in combination with another antiplatelet drug (without coprescription of an ulcer-healing drug) | Patients aged ≥18 years prescribed aspirin without coprescription of ulcer-healing drug (PPI or H2 antagonist) within the 3 months leading up to the audit date | Patients in the group denominator AND prescribed another antiplatelet drug (clopidogrel or prasugrel or ticagrelor) within the 3 months leading up to the audit date and within 28 days of the aspirin prescription |
| **Indicators associated with cautioned medication in other conditions (including heart failure, asthma and acute kidney injury)** |  |  |
| **HF & NSAID:** prescription of an oral NSAID to a patient with heart failure | Patients aged ≥18 years who have a diagnosis of heart failure at least 3 months before the audit date | Patients in the group denominator AND prescribed an oral NSAID within the 3 months leading up to the audit date |
| **Asthma & beta-blocker:** prescription of a non-selective beta-blocker to a patient with asthma | Patients aged ≥18 years with a Read code for asthma at least 3 months before audit date and no subsequent asthma resolved code during that time period | Patients in the group denominator AND prescribed a non-selective β-blocker within the 3 months leading up to the audit date |
| **CRF & NSAID:** prescription of an oral NSAID to a patient with eGFR <45 | Patients aged ≥18 years with chronic renal failure: eGFR <45 at least 3 months before the audit date | Patients in the group denominator AND prescribed an oral NSAID within the 3 months leading up to the audit date |
| **Indicators associated blood test monitoring** |  |  |
| **ACEI or loop diuretic, no blood tests:** patients aged 75 years and older who have been prescribed an angiotensin converting enzyme (ACE) inhibitor or a loop diuretic long term who have not had a computer-recorded check of their renal function and electrolytes in the previous 15 months | Patients aged ≥75 years prescribed an ACE inhibitor or a loop diuretic long-term i.e. first prescription for an ACE inhibitor or a loop diuretic at least 15 months prior to the audit date and at least one prescription (for the same drug) in the 6 months leading up to the audit date | Patients in the group denominator AND who have not had a computer-recorded check of their renal function and electrolytes within the previous 15 months leading up to the audit date |
| Patients receiving methotrexate for at least 3 months who have not had a recorded:<br>Full blood count (FBC) within the previous 3 months (**Methotrexate and no FBC**)<br>Liver function test (LFT) within the previous 3 months (**Methotrexate and no LFT**) | Patients aged ≥18 years with one or more prescriptions for methotrexate 3 to 6 months prior to the audit date and in the 3 months leading up to the audit date | Patients in the group denominator AND who have not had a computer-recorded:<br>FBC within the 3 months leading up to the audit date<br>LFT within the 3 months leading up to the audit date |
| **Lithium and no level recording:** patients receiving lithium for at least 3 months who have not had a recorded check of their lithium concentrations in the previous 3 months | Patients aged ≥18 years with one or more prescriptions for lithium recorded on computer 3 to 6 months prior to the audit date and in the 3 months leading up to the audit date | Patients in the group denominator AND who have not had a computer-recorded lithium level within the 3 months leading up to the audit date |
| **Amiodarone and no TFT:** patients receiving amiodarone for at least 6 months who have not had a thyroid function test (TFT) within the previous 6 months | Patients aged ≥18 years with one or more prescriptions for amiodarone 6 to 12 months prior to the audit date and in the 6 months leading up to the audit date | Patients in the group denominator AND who have not had a computer-recorded TFT within the 6 months leading up to the audit date |

Each indicator was specified in analytic code using PRIMIS SNOMED-CT codelists using the OpenSAFELY framework. We generated the numerator and denominator for each indicator per month between September 2019 and September 2021, and then calculated monthly percentages for each practice. For indicators assessing numeric values, only unambiguous results were used in the calculation of indicator percentages (for example, an eGFR value of “>30” was considered ambiguous for an indicator requiring the identification of patients with an eGFR value less than 45); note that this functionality was not available in OpenSAFELY-EMIS at the time of the study, so results for the *CRF & NSAID* indicator are reported for OpenSAFELY-TPP practices only.

The monthly indicator percentages were summarised as deciles and presented as decile charts across all practices each month. We also calculated the mean rate across practices in Q1 2020 and 2021 for each indicator as well as total counts of the numerator and denominator for each indicator across the two year period; note that in this cumulative data, repeated events will be counted for each month the event occurs (e.g., if a heart failure patient is prescribed an oral NSAID in two separate months (*HF & NSAID*), this is represented as two separate events). Across this period we also calculated the ratio of hazardous prescribing events to unique patients experiencing those events (to give an indication of the extent of repeated hazardous prescribing) and the number and percentage of practices with at least one instance of potentially hazardous prescribing at any point across the period.

Each blood test monitoring indicator has an associated monitoring window (e.g. lithium concentrations are required to be checked within 3 months). We annotated each decile plot to indicate this monitoring window, as calculated from the onset of COVID-19 in March 2020, for each indicator. We can interpret these lines as the point at which *all* patients would be subject to delayed blood test monitoring (a ‘worst case scenario’), should no action have been taken to rectify any COVID-19 related delays.

### Software and Reproducibility

Data management and analysis was performed using the OpenSAFELY software libraries and Python, both implemented using Python 3.8. A federated analysis involves carrying out patient-level analysis in multiple secure datasets, then later combining them: codelists and code for data management and data analysis were specified once using the OpenSAFELY tools; then transmitted securely to the OpenSAFELY-TPP platform within TPP’s secure environment, and separately to the OpenSAFELY-EMIS platform within EMIS’s secure environment, where they were each executed separately against local patient data; summary results were then reviewed for disclosiveness, released, and combined for the final outputs. All code for the OpenSAFELY platform for data management, analysis and secure code execution is shared for review and re-use under open licences at github.com/OpenSAFELY. Decile charts were drawn using Seaborn and matplotlib.

All analytical code used in this study is openly available for inspection and re-use at https://github.com/opensafely/pincer-measures. All codelists used are openly available at OpenSAFELY Codelists (https://www.opencodelists.org/codelist/pincer/). Results are available as an online report https://reports.opensafely.org/reports/changes-in-pincer-measures-throughout-the-covid-19-pandemic/.

### Patient and Public Involvement

This analysis relies on the use of large volumes of patient data. Ensuring patient, professional and public trust is therefore of critical importance. Maintaining trust requires being transparent about the way OpenSAFELY works, and ensuring patient voices are represented in the design of research, analysis of the findings, and considering the implications. For transparency purposes we have developed a public website (https://opensafely.org/) which provides a detailed description of the platform in language suitable for a lay audience; we have participated in two citizen juries exploring public trust in OpenSAFELY (13); we are currently co-developing an explainer video; we have ‘expert by experience’ patient representation on our OpenSAFELY Oversight Board; we have partnered with Understanding Patient Data to produce lay explainers on the importance of large datasets for research; we have presented at a number of online public engagement events to key communities; and more. To ensure the patient voice is represented, we are working closely with appropriate medical research charities.

## Results

We identified 1,813,058 patients registered across 6,367 practices at risk of potentially hazardous prescribing as indicated by the PINCER indicators at any point between September 2019 and September 2021. Demographic characteristics for 14,284,444 patients who were identified in at least one indicator denominator in the last month of the study period (September 2021) are provided in Table 2.

**Table 2.** Cohort description for any patients included in the denominator of at least one of the PINCER indicators at the end of the study period (September 2021), in OpenSAFELY-TPP and OpenSAFELY-EMIS. IMD=index of multiple deprivation.

| Characteristic | Category | OpenSAFELY-TPP |  | OpenSAFELY-EMIS |  | Total |  |
| --- | --- | --- | --- | --- | --- | --- | --- |
|  |  | n | % | n | % | n | % |
| Total |  | 5,998,805 | 100.00 | 8,285,639 | 100.00 | 14,284,444 | 100.00 |
| Age | 18-19 | 69,947 | 1.17 | 93,636 | 1.13 | 163,583 | 1.15 |
|  | 20-29 | 473,181 | 7.89 | 679,780 | 8.20 | 1,152,961 | 8.07 |
|  | 30-39 | 507,839 | 8.47 | 788,895 | 9.52 | 1,296,734 | 9.08 |
|  | 40-49 | 468,532 | 7.81 | 689,394 | 8.32 | 1,157,926 | 8.11 |
|  | 50-59 | 527,714 | 8.80 | 766,127 | 9.25 | 1,293,841 | 9.06 |
|  | 60-69 | 1,217,103 | 20.29 | 1,647,347 | 19.88 | 2,864,450 | 20.05 |
|  | 70-79 | 1,672,710 | 27.88 | 2,235,918 | 26.99 | 3,908,628 | 27.36 |
|  | 80+ | 1,061,779 | 17.70 | 1,384,542 | 16.71 | 2,446,321 | 17.13 |
| Sex | F | 3,122,219 | 52.05 | 4,291,110 | 51.79 | 7,413,329 | 51.90 |
|  | M | 2,876,586 | 47.95 | 3,994,529 | 48.21 | 6,871,115 | 48.10 |
| Ethnicity | White | 4,285,420 | 71.44 | 5,480,579 | 66.15 | 9,765,999 | 68.37 |
|  | South Asian | 209,386 | 3.49 | 405,580 | 4.89 | 614,966 | 4.31 |
|  | Black | 69,225 | 1.15 | 199,570 | 2.41 | 268,795 | 1.88 |
|  | Other | 48,206 | 0.80 | 77,498 | 0.94 | 125,704 | 0.88 |
|  | Mixed | 38,392 | 0.64 | 84,486 | 1.02 | 122,878 | 0.86 |
|  | Missing | 1,348,176 | 22.47 | 2,037,926 | 24.60 | 3,386,102 | 23.70 |
| IMD | Most deprived (1) | 984,981 | 16.42 | 1,425,367 | 17.20 | 2,410,348 | 16.87 |
|  | 2 | 1,094,257 | 18.24 | 1,569,287 | 18.94 | 2,663,544 | 18.65 |
|  | 3 | 1,306,389 | 21.78 | 1,649,173 | 19.90 | 2,955,562 | 20.69 |
|  | 4 | 1,283,629 | 21.40 | 1,741,211 | 21.01 | 3,024,840 | 21.18 |
|  | Least deprived (5) | 1,227,902 | 20.47 | 1,875,295 | 22.63 | 3,103,197 | 21.72 |
|  | Missing | 101,647 | 1.69 | 25,306 | 0.31 | 126,953 | 0.89 |
| Region | East | 1,337,146 | 22.29 | 343,151 | 4.14 | 1,680,297 | 11.76 |
|  | London | 276,220 | 4.60 | 1,431,617 | 17.28 | 1,707,837 | 11.96 |
|  | Midlands<sup>1</sup> | 1,282,238 | 21.37 | 1,494,269 | 18.03 | 2,776,507 | 19.44 |
|  | North East & Yorkshire<sup>2</sup> | 1,548,534 | 25.81 | 702,833 | 8.48 | 2,251,367 | 15.76 |
|  | North West | 94,912 | 1.58 | 1,693,016 | 20.43 | 1,787,928 | 12.52 |
|  | South East | 502,990 | 8.38 | 1,905,796 | 23.00 | 2,408,786 | 16.86 |
|  | South West | 956,765 | 15.95 | 714,957 | 8.63 | 1,671,722 | 11.70 |

<sup>1</sup> Comprised of *East Midlands* and *West Midlands* in OpenSAFELY-TPP
<sup>2</sup> Comprised of *Yorkshire and the Humber* and *North East* in OpenSAFELY-TPP

For each PINCER indicator, we show Q1 mean percentages for 2020 and 2021 to enable comparison between a pre-COVID-onset and post-COVID-onset point in time (Table 3). Mean Q1 2020 percentages ranged from 1.11% (*Age ≥ 65 & NSAID*) to 36.20% (*Amiodarone and no TFT*), while Q1 2021 percentages ranged from 0.75% (*Age ≥ 65 & NSAID*) to 39.23% (*Amiodarone and no TFT*). The pre-/post-COVID-onset difference ranged from a reduction of 0.59% (*Warfarin/DOAC & antiplatelet*) to an increase of 6.98% (*ACEI or loop diuretic, no blood tests*).

**Table 3.**
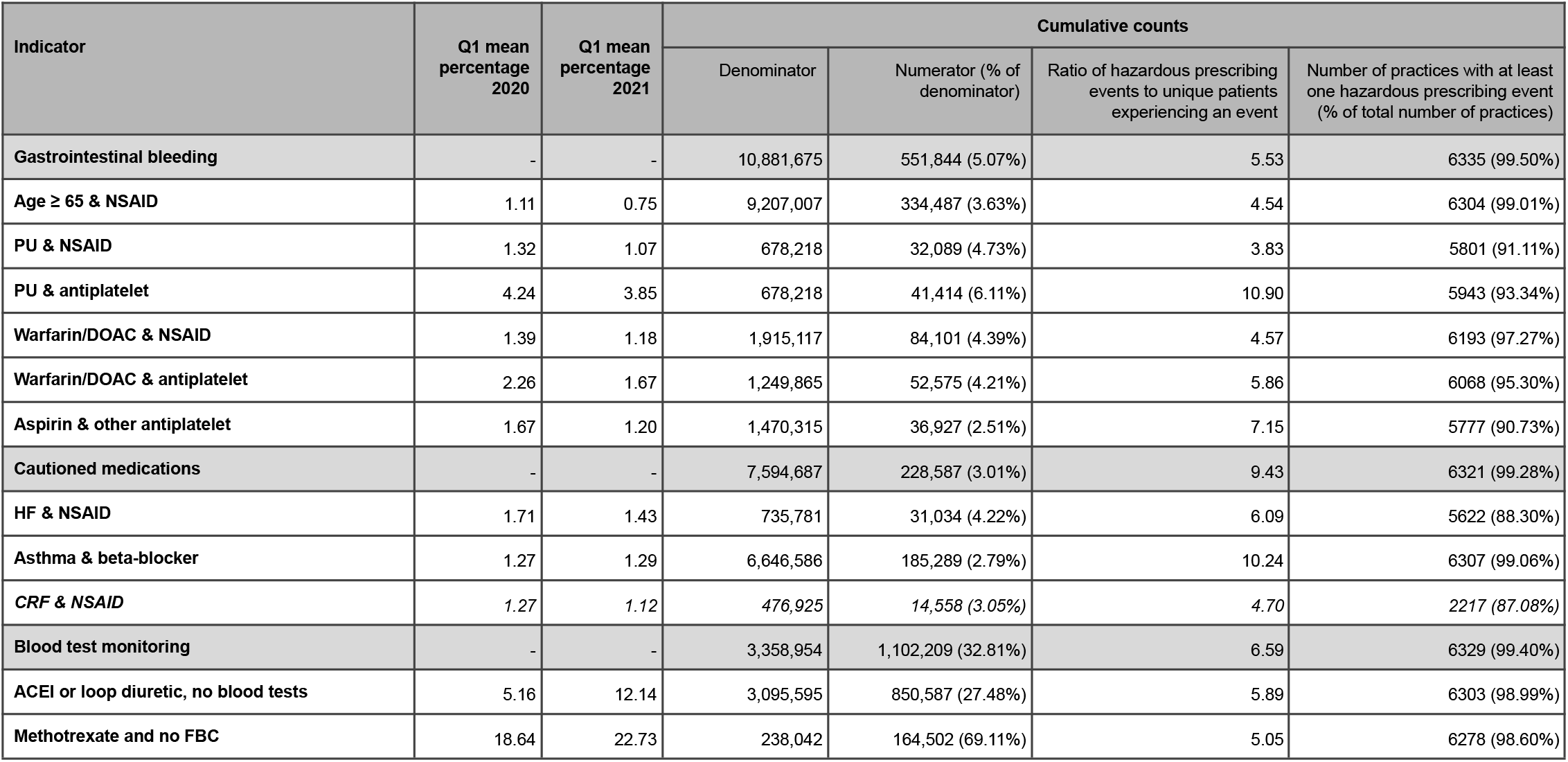
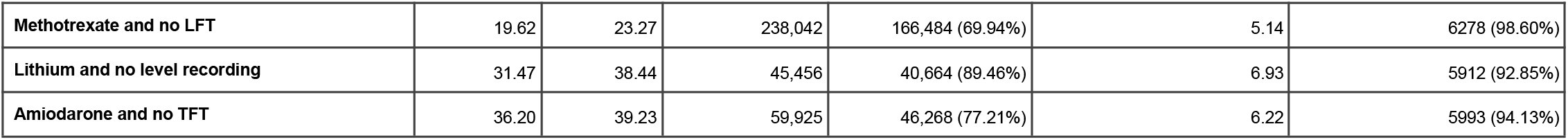
Indicator rates for PINCER hazardous prescribing indicators: Q1 2020/2021 percentages and cumulative results between September 2019 and September 2021. Mean values are calculated at the practice-level. Rates for CRF & NSAID (italicised) are calculated across 2546 OpenSAFELY-TPP practices; all other indicator rates are calculated across all 6367 practices (2546 OpenSAFELY-TPP + 3821 OpenSAFELY-EMIS practices). Tables for OpenSAFELY-TPP and OpenSAFELY-EMIS separately are available in Supplementary Tables 1-2.

Cumulative counts for each indicator are provided in Table 3. The percentage of patients identified as at risk of a potentially hazardous prescribing event in the study period ranged from 2.51% (36,927 of 1,470,315 patients; *Aspirin & other antiplatelet*) to 89.46% (40,664 of 45,456 patients; *Lithium and no level recording*). The ratio of hazardous events to patients ranged from 3.83 (*PU & NSAID*) to 10.90 (*PU & antiplatelet*). The percentage of practices with an event for each indicator ranged from 87.02% (*CRF & NSAID*) to 99.06% (*Asthma & beta-blocker*).

### Indicators associated with gastrointestinal bleeding

The six indicators of potentially hazardous prescribing in relation to GI bleeds exhibit a decreasing trend across the study period with minimal impact of the pandemic (Figure 1a, OpenSAFELY-TPP only and OpenSAFELY-EMIS only decile charts are provided in Supplementary Figures 1a and 2a respectively). The mean percentage of *Warfarin/DOAC & NSAID* was 1.39% in Q1 2020; the equivalent rate in 2021 was 1.18%. Similarly, the percentage of *PU & antiplatelet* fell from 4.24% to 3.85% between Q1 2020 and Q1 2021 (Table 3). There were only marginal impacts on this improving trend during the peak waves of the COVID-19 pandemic.

**Figure 1.**
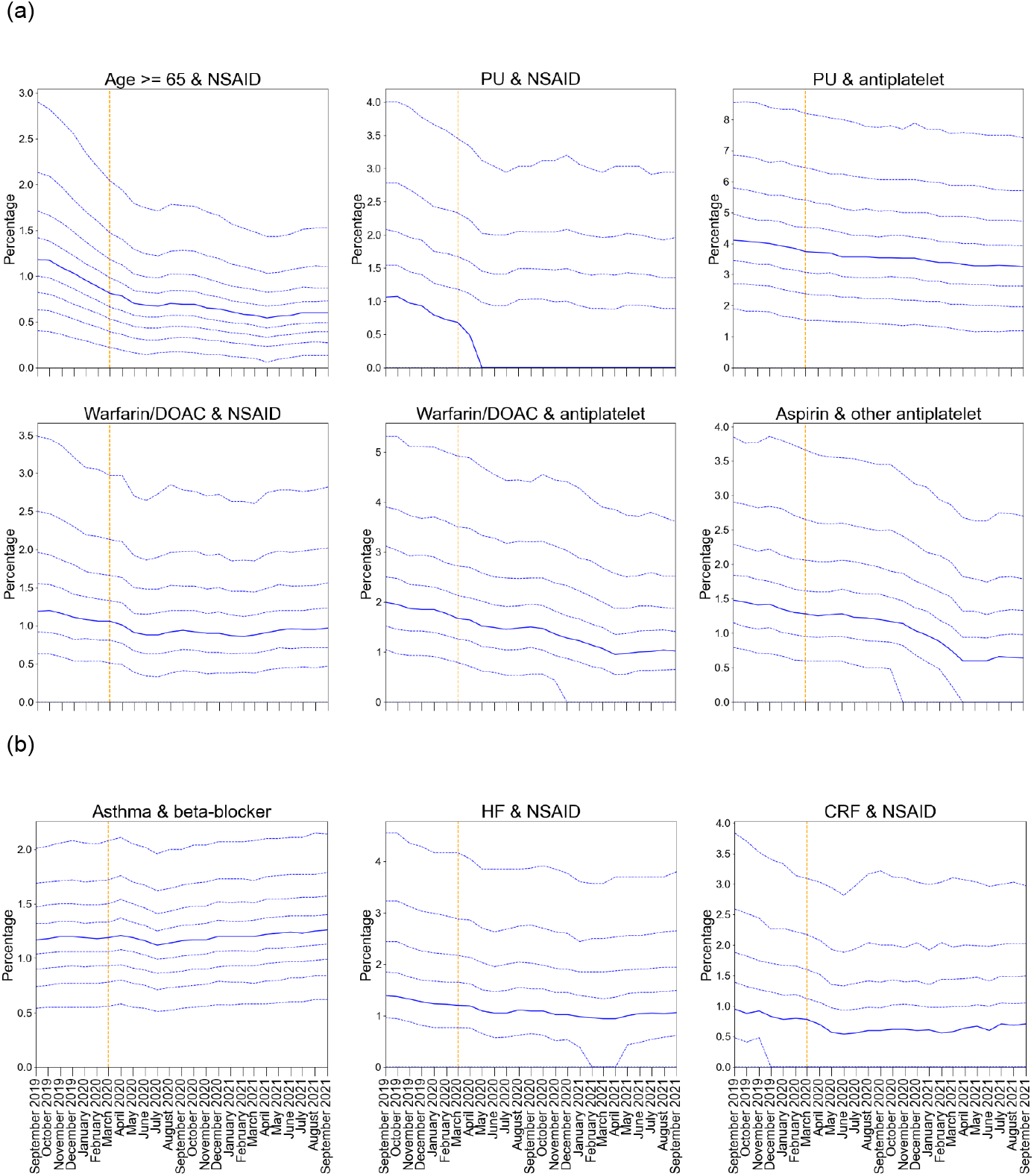
Practice level decile plots for PINCER prescribing indicators, specifically in relation to (a) GI bleeding and (b) cautioned medications. The percentage of patients identified as at risk of potentially hazardous prescribing as measured by each indicator is reported for the period September 2019 to September 2021 (inclusive). The median percentage is displayed as a thick blue line and deciles are indicated by dashed blue lines. The month of national lockdown in England as a response to the onset of COVID-19 (March 2020) is highlighted with an orange dashed vertical line. Deciles for CRF & NSAID are calculated across 2546 OpenSAFELY-TPP practices; all other deciles are calculated across 6367 practices (2546 OpenSAFELY-TPP + 3821 OpenSAFELY-EMIS practices).

### Indicators associated with cautioned medications

Time trends and variation for all three “cautioned medication” indicators are presented in Figure 1b (OpenSAFELY-TPP only and OpenSAFELY-EMIS only decile charts are provided in Supplementary Figures 1b and 2b respectively): COVID-19 had minimal impact on compliance for all indicators.

### Indicators associated with blood test monitoring

All blood test monitoring indicators exhibited an increase in delayed monitoring immediately following the onset of COVID-19 (May-July 2020; Figure 2, OpenSAFELY-TPP only and OpenSAFELY-EMIS only decile plots are provided in Supplementary Figures 3-4 respectively).These increased rates showed considerable recovery by August-September 2020, in the case of *Lithium and no level recording* (31.47% Q1 2020 vs 38.44% Q1 2021), *Methotrexate and no FBC* (18.64% Q1 2020 vs 22.73% Q1 2021) and *Methotrexate and no LFT* (19.62% Q1 2020 vs 23.27% Q1 2021). The *Amiodarone and no TFT* indicator behaved similarly (36.20% Q1 2020 vs 39.23% Q1 2021), though the initial post-COVID-onset recovery period extended into October-November 2020 (Table 3).

**Figure 2.**
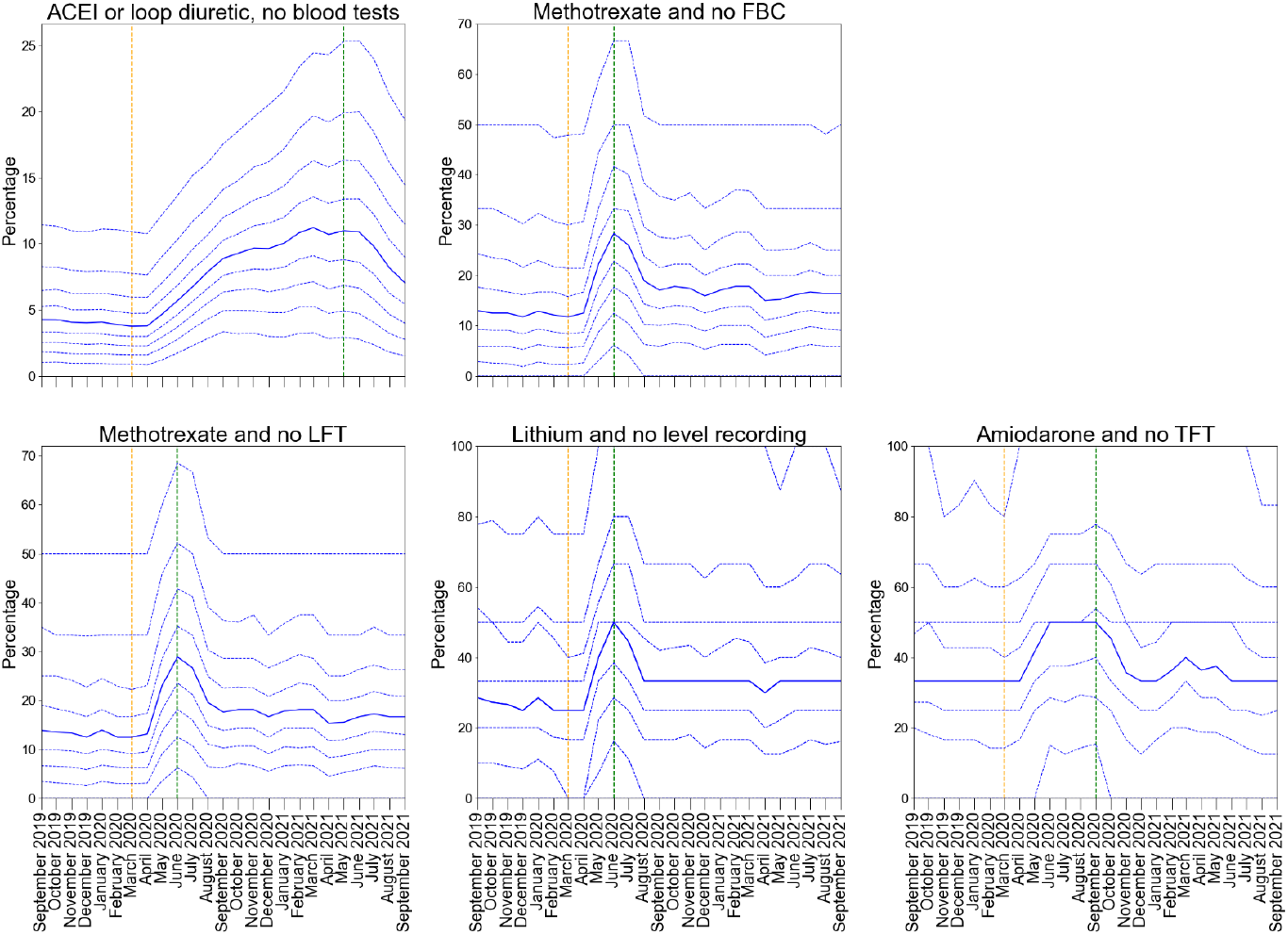
Practice level decile plots for PINCER blood test monitoring indicators. The percentage of patients identified as at risk of potentially blood test monitoring as measured by each indicator is reported for the period September 2019 to September 2021 (inclusive). The median percentage is displayed as a thick blue line and deciles are indicated by dashed blue lines. The month of national lockdown in England as a response to the onset of COVID-19 (March 2020) is highlighted with an orange dashed vertical line. The monitoring window, as measured from the onset of COVID-19, for each indicator is shown by a green dashed vertical line. All deciles are calculated across 6367 practices (2546 OpenSAFELY-TPP + 3821 OpenSAFELY-EMIS practices).

The “*ACEI or loop diuretic, no blood tests”* indicator exhibited a noticeably different COVID-19 response pattern than the other blood test monitoring indicators. Here, the monitoring worsened steadily over a longer period of time, increasing from a mean of 5.16% to 12.14% between Q1 2020 and Q1 2021, beginning to recover in June 2021. The assessment window for this indicator is significantly wider than the windows for the other blood test monitoring indicators: within 15 months of prescription compared to three (lithium and methotrexate) or six (amiodarone) months.

## Discussion

### Summary

Despite substantial barriers to the delivery of primary care during the COVID-19 pandemic, good performance was maintained across a diverse range of widely evaluated and nationally adopted measures of safe prescribing. There were evident delays in delivering some medication-related blood test monitoring within the time-window specified in the safety measure; especially for those blood tests where the time-window for compliance is itself already very long, and tests infrequent. However all indicators exhibited considerable recovery by the end of the study period. It was possible to describe PINCER indicators for almost all patients and GP practices in England with a single analysis in OpenSAFELY.

### Strengths and limitations

This study has a range of strengths, some unprecedented. The scale and completeness of the data in the OpenSAFELY platform is greater than that through any other route for accessing GP data: other approaches either offer substantially fewer active current patients, in a data download onto researchers’ local machine (such as Clinical Practice Research Datalink); or only contain a small derived subset of information in the GP record (such as the GPES dataset extracted and disseminated by NHS Digital). Previous audits for compliance with PINCER or similar measures and indicators in primary care rely on manual audit within a practice, or analyses on data downloaded from a group of practices. By contrast OpenSAFELY executes analyses in a secure environment inside the EHR provider data centre, across the full set of all structured data in the GP record including all tests, prescriptions, diagnostic codes and referrals. In addition, although the underlying GP data is stored in two very different settings (TPP and EMIS) analyses written once in OpenSAFELY then execute in each setting identically, with the outputs aggregated afterwards, in a process known as “federated analytics”. Overall this represents a unique, national platform able to capture the patient journey for 57 million people in England whilst prioritising patient privacy.

A second strength is the transparency and reproducibility of the analysis: as with all OpenSAFELY analyses, the complete set of all code for the platform and for all data curation and analysis for every study from raw data to completed output is shared openly on GitHub in standard formats for scientific review and efficient re-use under open licences by all.

A third strength is the robustness of the measures for each safety behaviour: all eligible patients and targeted clinical safety behaviours were developed for the national PINCER medication safety programme which has been extensively peer reviewed and evaluated throughout the NHS over many years with strong support from clinicians and commissioners.

We also note some limitations. We note that our data will only include test results carried out in primary care, or those in secondary care that are returned to GPs as structured data: this may therefore not include test results communicated by letter or phone (such as tests requested while a person is in hospital, or other settings like psychiatric outpatients). However this is in line with the methodology already used in the national PINCER programme to evaluate compliance with the targeted safety behaviours using primary care data alone.

### Comparison of existing literature

A recent systematic review of healthcare usage during the pandemic, encompassing 81 studies across 20 countries found that healthcare utilisation (including visits, admissions, diagnostics and therapeutics) reduced by 37% during the pandemic, highlighting a substantial reduction in April-May 2020 (14). The WHO also identified significant disruption to countries’ healthcare capacity for non-communicable diseases in a rapid assessment in May 2020 (15). A population cohort-based study conducted using the OpenSAFELY platform reported that clinical activity in relation to blood tests declined in the months following COVID-19 onset, but also reported recovery of these same tests by September 2020 (16). These findings are in line with our observations of the blood test monitoring indicators, where substantial delays were experienced in the same period of time. Interrupted service delivery leading to reduced NSAID prescriptions following acute presentations may also explain the temporary reduction of the first GI bleed indicator (*prescription of an oral NSAID, without co-prescription of an ulcer-healing drug, to a patient aged ≥65 years*) in April-July 2020; this is supported by data for this period in OpenPrescribing (17) and lower than predicted rates of prescribing for Naproxen and Ibuprofen in this period (18). Elsewhere we have found evidence of prioritisation of anticoagulant services, with blood tests to manage high-risk anticoagulants being prioritised during the initial stages of the COVID-19 pandemic (16); data from the current study also suggests that prescribing in relation to anticoagulants is a priority, with all GI bleed indicators being unaffected, and continuing to decline, following COVID-19 onset.

In the early stages of the pandemic, in recognition of the increased risk of medication related harm during the COVID-19 pandemic, NHS England and local Clinical Commissioning Groups (CCGs) revised guidance regarding blood test monitoring, extending the recommended monitoring window for some patient populations (e.g., in relation to lithium (19) and methotrexate (20)) or advising that blood monitoring for lower risk medications should be carried out “if possible” (e.g., ACE inhibitors (21)), if clinically safe to do so. There is some evidence in our data that practices did adopt this revised guidance, with post-recovery blood test monitoring often falling just short of pre-pandemic levels (particularly in the case of methotrexate and lithium).

### Implications for policy and research

The variation in service recovery observed in the blood test monitoring indicators may in part be due to the assessment window for each indicator, and clinicians prioritising urgent work during the pandemic. For example, it is possible that the protracted recovery of the ACE inhibitor monitoring was due to primary care services proactively prioritising monitoring of higher risk prescriptions such as methotrexate so as to minimise the impact of service disruption on patient care. It is also likely that the systems put in place around the monitoring of high risk drugs (e.g., clinical system alerts) contributed towards expedited recovery of the other blood test monitoring indicators, particularly in the case of lithium and methotrexate. The decile chart for this indicator starts to plateau well before the ‘worst case scenario’ timepoint, suggesting that the majority of primary care providers successfully implemented recovery programmes in this clinical domain. Further areas for research include using innovative change detection methodologies (22) to ascertain practice-level features that influence recovery and resilience in the context of service disruption to inform WHO and NHS England’s recommendations to “build back better”.

The implications of this analysis for data usage in the NHS are very substantial. Historically, due to practical and privacy challenges around accessing GP data at scale, each practice participating in the PINCER programme has been required to manually execute the necessary computerised searches before individually uploading their results for central oversight; in some centres data for a group of practices and patients can be downloaded and analysed in larger volumes. This manual approach introduces delays and increases the resource cost of monitoring safety. Using the OpenSAFELY framework we were able to execute a single analysis for almost the entire population of England in near-real-time, while leaving data in situ. This approach is efficient: analyses can be easily updated, and expanded, because they are executed in a single framework from re-executable code. It also preserves patient trust: OpenSAFELY was the single most highly trusted COVID-19 data project in a rigorous Citizens Jury sponsored by the NHS and the National Data Guardian (13). Furthermore, the additional data also securely accessible through the OpenSAFELY tools can be used to describe PINCER measures in fine-grained demographic or clinical sub-populations. These tools can facilitate near real-time audit and feedback in the context of rapidly evolving pressures on the health service and are readily extendable to other clinical and challenges.

More broadly this analysis demonstrates the power of collaborative working with shared open source code in the NHS: it built upon the work of PRIMIS in establishing and then publicly releasing the full code for a set of rigorously tested medication safety indicators; and then implemented the open code from PINCER in the open source framework of OpenSAFELY, to assess a critical public health question on a national scale. Open working as demonstrated here is strongly supported by senior stakeholders in multiple sectors (23–25) and can bring substantial benefits: it facilitates efficient re-use of prior technical work; it ensures fidelity through the consistent implementation of data curation and analysis across all organisations; it supports complete reproducibility; it enables error-checking by all interested parties; and it facilitates public and professional trust.

### Conclusion

NHS GP data can be analysed at national scale to generate insights on service delivery with strong public support. Potentially hazardous prescribing was largely unaffected by COVID-19 in a dataset of 57 million patients’ full primary care health records in England.

## Supporting information

Supplementary Tables 1-2 (legends included)

Supplementary Figures 1-4 (legends included)

STROBE checklist

## Data Availability

https://www.opencodelists.org/codelist/pincer/

https://reports.opensafely.org/reports/changes-in-pincer-measures-throughout-the-covid-19-pandemic/

## Acknowledgements

We are very grateful for all the support received from the EMIS and TPP Technical Operations team throughout this work, and for generous assistance from the information governance and database teams at NHS England / NHSX.

## Administrative

### Funding

This work was jointly funded by UKRI, NIHR and Asthma UK-BLF [COV0076; MR/V015737/] and the Longitudinal Health and Wellbeing strand of the National Core Studies programme. The OpenSAFELY data science platform is funded by the Wellcome Trust.

B.G.’s work on better use of data in healthcare more broadly is currently funded in part by: the Wellcome Trust, NIHR Oxford Biomedical Research Centre, NIHR Applied Research Collaboration Oxford and Thames Valley, the Mohn-Westlake Foundation; all DataLab staff are supported by B.G.’s grants on this work. B.M.K. is also employed by NHS England working on medicines policy and clinical lead for primary care medicines data. The views expressed are those of the authors and not necessarily those of the NIHR, NHS England, Public Health England or the Department of Health and Social Care.

The underlying research, and the development and rollout out of the PINCER intervention was funded by the Department of Health Patient Safety Research Portfolio (original PINCER trial); the Health Foundation and Academic Health Science Network East Midlands (rollout across the East Midlands as part of the Health Foundation *Scaling Up Improvement* Programme), and NHS England, the national Academic Health Science Network and the Health Foundation (national rollout of PINCER). A.J.A.’s current work on the evaluation of the PINCER intervention is funded by NIHR Programme Grants for Applied Research (RP-PG-1214-20012).

The views expressed are those of the authors and not necessarily those of the NIHR, NHS England, Public Health England or the Department of Health and Social Care.

Funders had no role in the study design, collection, analysis, and interpretation of data; in the writing of the report; and in the decision to submit the article for publication.

### Author contributions

**Conceptualization:** L.F., L.E.M.H., K.O., S.R., D. Evans, B.G., and B.M.

**Data curation:** K.O., S.R., J.B., D. Evans, A.M., S.B., S.D., I.D., D. Evans, G.H., P.I., C.E.M., B.S., T.W., C.B., J.C., J.P., F.H., S.H., S.O.H., A.E., R.J., D.A., P.G., A.F., and N.P.

**Formal analysis:** L.F., L.E.M.H., and B.M.

**Funding acquisition:** K.O., S.R., A.J.A., D. Evans, J. Morley, and B.G.

**Investigation:** L.F., L.E.M.H., K.O., S.R., J.B., A.J.A., D. Evans, H.C., R.C., and B.M.

**Methodology:** L.F., L.E.M.H., and B.M.

**Resources:** K.O., S.R., J.B., A.J.A., D. Evans, A.M., S.B., S.D., I.D., D. Evans, G.H., P.I., C.E.M., B.S., T.W., C.B., J.C., J.P., F.H., S.H., S.O.H., A.E., R.J., D.A., P.G., A.F., and N.P.

**Software:** S.B., S.D., I.D., D. Evans, G.H., P.I., C.E.M., B.S., T.W., C.B., and J.C.

**Supervision:** B.G.

**Visualisation:** L.F., L.E.M.H., and B.M.K

**Writing - original draft:** L.F., L.E.M.H., H.C., and B.M.K.

**Writing - review & editing:** L.F., L.E.M.H., K.O., S.R., J.B., A.J.A., D. Evans, H.C., R.C., O.M., J. Morley, A.M., W.H., A.G., J. Massey, A.J.W., B.G., and B.M.

### Information governance

NHS England is the data controller; EMIS and TPP are the data processors and the key researchers on OpenSAFELY are acting with the approval of NHS England. This implementation of OpenSAFELY is hosted within the EMIS and TPP environments which are accredited to the ISO 27001 information security standard and are NHS IG Toolkit compliant;(26,27) patient data has been pseudonymised for analysis and linkage using industry standard cryptographic hashing techniques; all pseudonymised datasets transmitted for linkage onto OpenSAFELY are encrypted; access to the platform is via a virtual private network (VPN) connection, restricted to a small group of researchers; the researchers hold contracts with NHS England and only access the platform to initiate database queries and statistical models; all database activity is logged; only aggregate statistical outputs leave the platform environment following best practice for anonymisation of results such as statistical disclosure control for low cell counts.(28) The OpenSAFELY research platform adheres to the obligations of the UK General Data Protection Regulation (GDPR) and the Data Protection Act 2018. In March 2020, the Secretary of State for Health and Social Care used powers under the UK Health Service (Control of Patient Information) Regulations 2002 (COPI) to require organisations to process confidential patient information for the purposes of protecting public health, providing healthcare services to the public and monitoring and managing the COVID-19 outbreak and incidents of exposure; this sets aside the requirement for patient consent.(29) Taken together, these provide the legal bases to link patient datasets on the OpenSAFELY platform. GP practices, from which the primary care data are obtained, are required to share relevant health information to support the public health response to the pandemic, and have been informed of the OpenSAFELY analytics platform.

The lead author (B.G.) affirm that the manuscript is an honest, accurate, and transparent account of the study being reported; that no important aspects of the study have been omitted; and that any discrepancies from the study as planned (and, if relevant, registered) have been explained.

### Ethical Approval

This study was approved by the Health Research Authority (REC reference 20/LO/0651) and by the LSHTM Ethics Board (reference 21863).

### Data sharing

All data were linked, stored and analysed securely within the OpenSAFELY platform (https://opensafely.org/). Data include pseudonymised data such as coded diagnoses, drugs, and physiological parameters. No free text data are included. All code is shared openly for review and reuse under MIT open license (https://github.com/opensafely/pincer-measures). Detailed pseudonymised patient data are potentially reidentifiable and therefore not shared.

### Dissemination to participants and related patient and public communities

We will share information and interpretation of our findings through press releases, social media channels, and plain language summary.

