## Supplementary Figures 1-4 (legends included) for "Changes in English medication safety indicators throughout the COVID-19 pandemic: a federated analysis of 57 million patients’ primary care records in situ using OpenSAFELY"

#### Supplementary Figure 1: GI bleed + cautioned (TPP only)

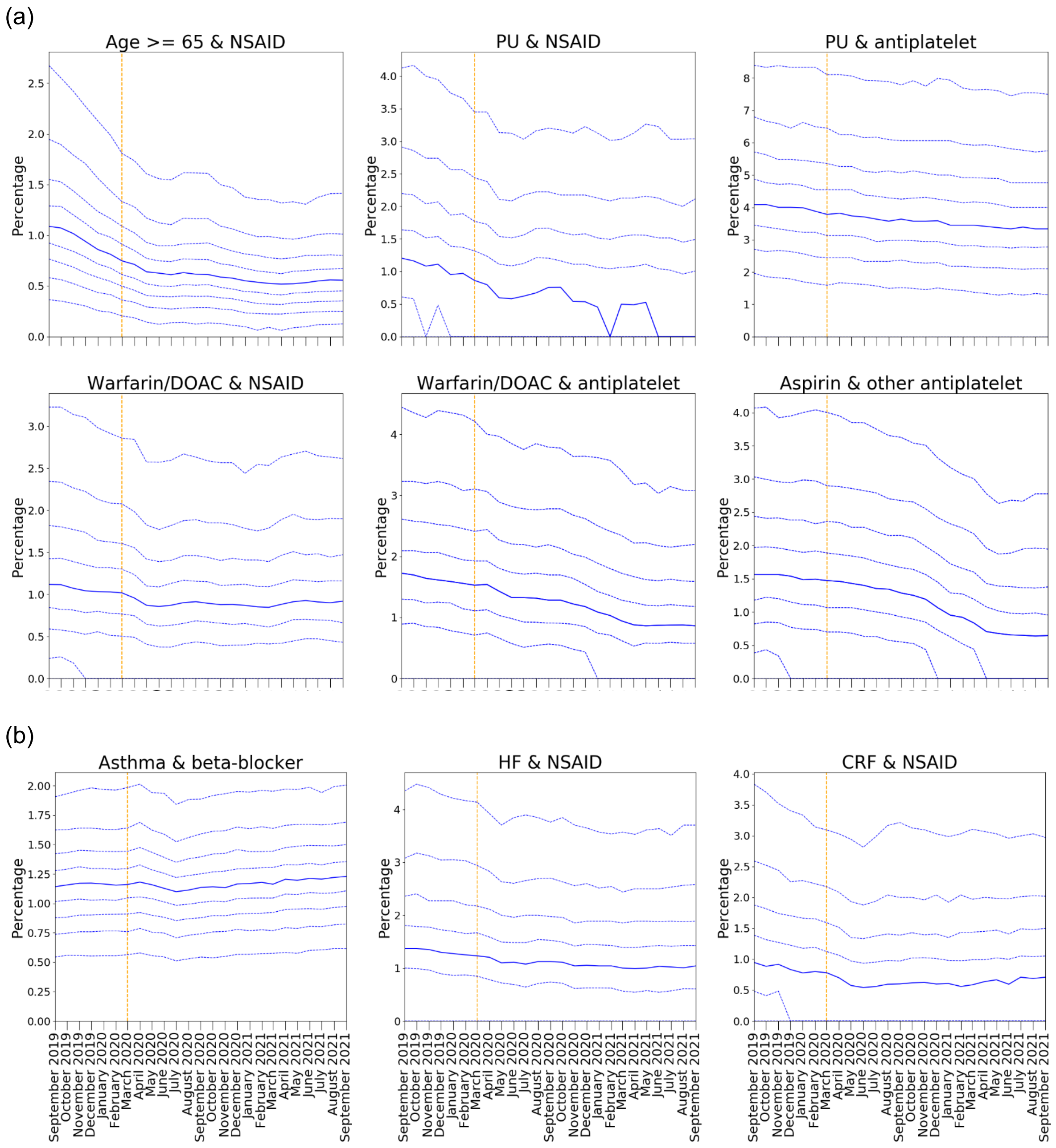


**Supplementary Figure 1 - OpenSAFELY-TPP practice level decile plots for PINCER prescribing indicators, specifically in relation to (a) GI bleeding and (b) cautioned medications.** All deciles are calculated across 2546 OpenSAFELY-TPP practices. The percentage of patients identified as at risk of potentially hazardous prescribing as measured by each indicator is reported for the period September 2019 to September 2021 (inclusive). The median percentage is displayed as a thick blue line and deciles are indicated by dashed blue lines. The month of national lockdown in England as a response to the onset of COVID-19 (March 2020) is highlighted with an orange dashed vertical line.

#### Supplementary Figure 2: GI bleed + cautioned (EMIS only)

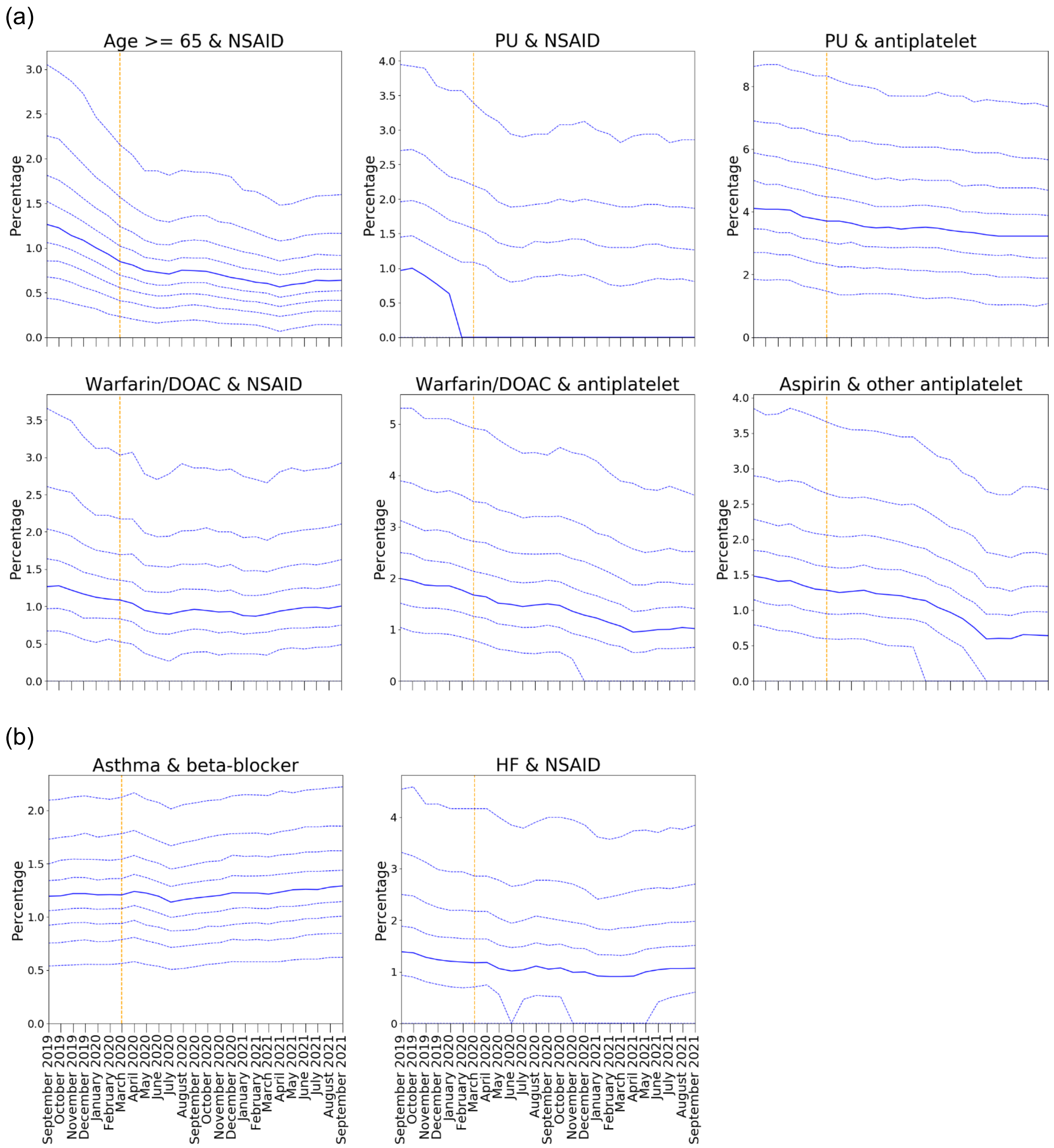


**Supplementary Figure 2 - OpenSAFELY-EMIS practice level decile plots for PINCER prescribing indicators, specifically in relation to (a) GI bleeding and (b) cautioned medications.** All deciles are calculated across 3821 OpenSAFELY-EMIS practices. The percentage of patients identified as at risk of potentially hazardous prescribing as measured by each indicator is reported for the period September 2019 to September 2021 (inclusive). The median percentage is displayed as a thick blue line and deciles are indicated by dashed blue lines. The month of national lockdown in England as a response to the onset of COVID-19 (March 2020) is highlighted with an orange dashed vertical line. Note that the CRF & NSAID indicator could not be implemented in OpenSAFELY-EMIS and therefore not shown.

### Supplementary Figure 3: monitoring (TPP only)

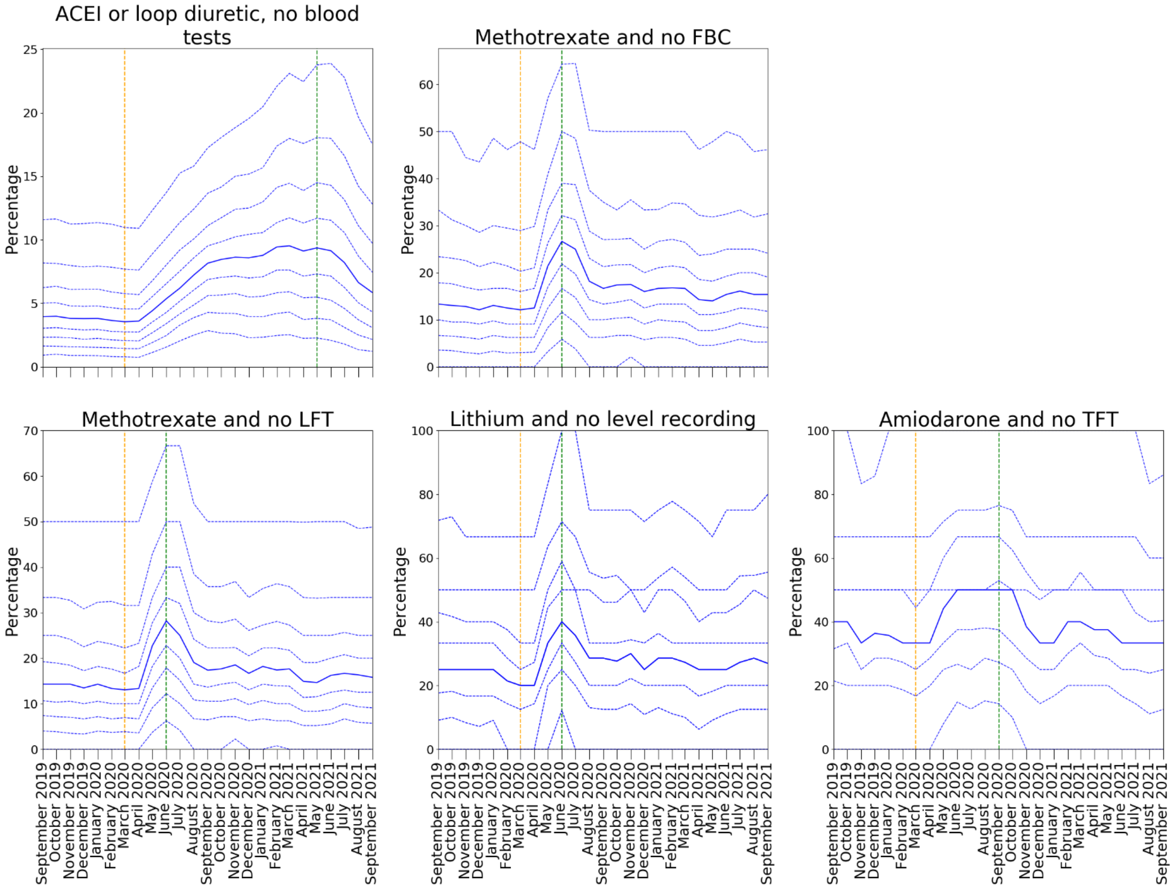


**Supplementary Figure 3 - OpenSAFELY-TPP practice level decile plots for PINCER blood test monitoring indicators.** All deciles are calculated across 2546 OpenSAFELY-TPP practices. The percentage of patients identified as at risk of potentially blood test monitoring as measured by each indicator is reported for the period September 2019 to September 2021 (inclusive). The median percentage is displayed as a thick blue line and deciles are indicated by dashed blue lines. The month of national lockdown in England as a response to the onset of COVID-19 (March 2020) is highlighted with an orange dashed vertical line. The monitoring window, as measured from the onset of COVID-19, for each indicator is shown by a green dashed vertical line.

### Supplementary Figure 4: monitoring (EMIS only)

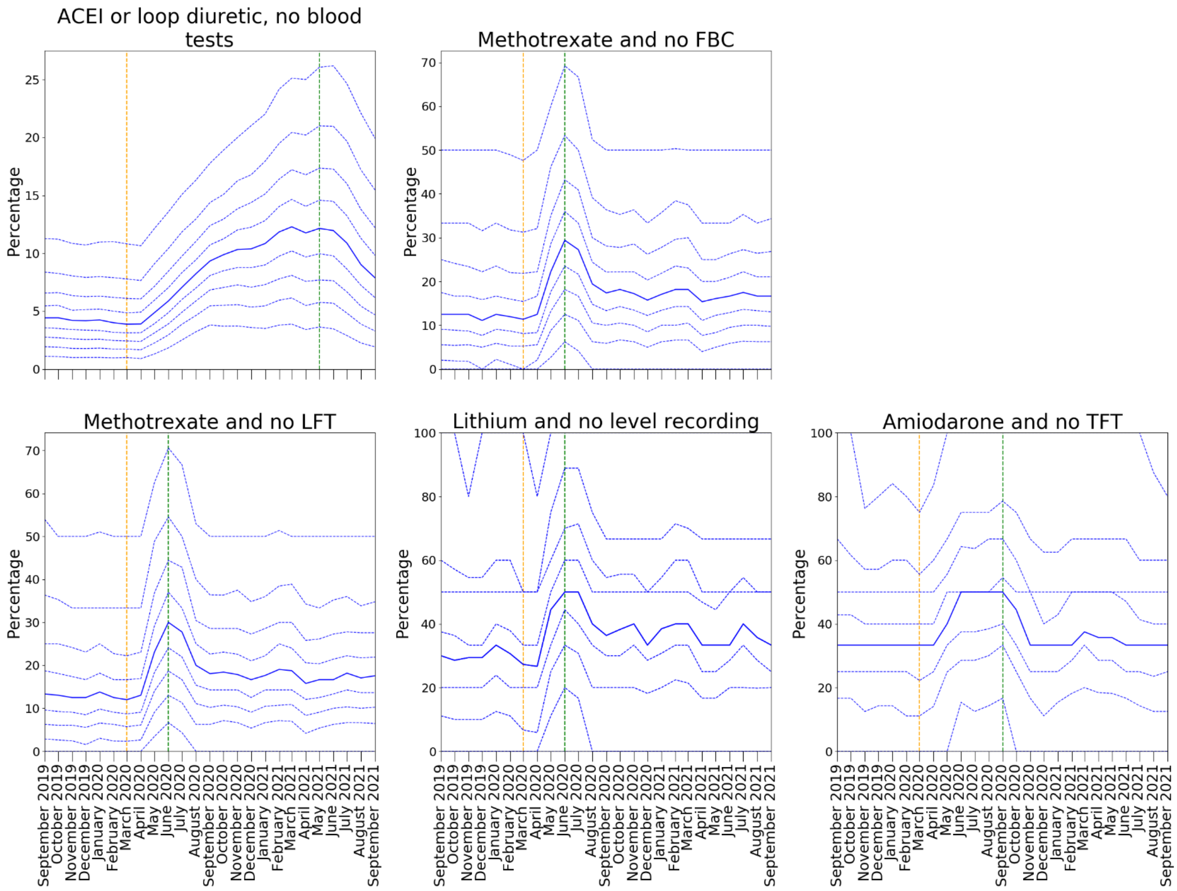


**Supplementary Figure 4 - OpenSAFELY-EMIS practice level decile plots for PINCER blood test monitoring indicators.** All deciles are calculated across 3821 OpenSAFELY-EMIS practices. The percentage of patients identified as at risk of potentially blood test monitoring as measured by each indicator is reported for the period September 2019 to September 2021 (inclusive). The median percentage is displayed as a thick blue line and deciles are indicated by dashed blue lines. The month of national lockdown in England as a response to the onset of COVID-19 (March 2020) is highlighted with an orange dashed vertical line. The monitoring window, as measured from the onset of COVID-19, for each indicator is shown by a green dashed vertical line.
